# A primary care approach to the COVID-19 pandemic: clinical features and natural history of 2,073 suspected cases in the Corona São Caetano programme, São Paulo, Brazil

**DOI:** 10.1101/2020.06.23.20138081

**Authors:** Fabio E Leal, Maria C Mendes-Correa, Lewis F Buss, Silvia F Costa, Joao CS Bizario, Sonia RP de Souza, Osorio Thomaz, Tania R Tozetto-Mendoza, Lucy S Villas-Boas, Lea CO Silva, Regina MZ Grespan, Ligia Capuani, Renata Buccheri, Helves Domingues, Neal DE Alexander, Philippe Mayaud, Ester C Sabino

## Abstract

**Background:** Despite most cases not requiring hospital care, there are limited community-based clinical data on COVID-19.

**Methods and findings:** The Corona São Caetano program is a primary care initiative offering COVID-19 care to all residents of São Caetano do Sul, Brazil. After triage of potentially severe cases, consecutive patients presenting between 13th April and 13th May 2020 were tested at home with SARS-CoV-2 reverse transcriptase (RT) PCR; positive patients were followed up for 14 days. RT-PCR-negative patients were offered SARS-CoV-2 serology. We describe the clinical features, virology and natural history of this prospective population-based cohort. Of 2,073 suspected COVID-19 cases, 1,583 (76·4%) were tested by RT-PCR, of whom 444 (28·0%, 95%CI: 25·9% - 30·3%) were positive; 604/1,136 (53%) RT-PCR-negative patients underwent serology, of whom 52 (8·6%) tested SARS-CoV-2 seropositive. The most common symptoms of COVID-19 were cough, fatigue, myalgia and headache; whereas self-reported fever, anosmia, and ageusia were most associated with a positive COVID-19 diagnosis. RT-PCR cycle thresholds were lower in men, older patients, those with fever and arthralgia, and around symptom onset. The rates of hospitalization and death among 444 RT-PCR-positive cases were 6·7% and 0·7%, respectively, with older age and obesity more frequent in the hospitalized group.

**Conclusions:** COVID-19 presents similarly to other mild respiratory disease in primary care. Some symptoms assist the differential diagnosis. Most patients can be managed at home.

## INTRODUCTION

A comprehensive public health response is vital but difficult to achieve during an epidemic. The COVID-19 pandemic, caused by the novel severe acute respiratory syndrome coronavirus 2 (SARS-CoV-2), started in China in late 2019.^1^ According to the World Health Organization (WHO)^2^ and others^3^, the ideal early response should have been multipronged, with identification, isolation, treatment and contact tracing of symptomatic cases, relying on a strong testing programme. Primary health care (PHC) is well placed to implement such a response, by identifying cases early and managing them in a way that minimizes overcrowding of emergency rooms and intensive care units.^4^ Real-time data analysis coming from these primary care response systems can inform policy decisions.

In Brazil, the first case of COVID-19 was identified in the city of São Paulo on 26th February 2020.^5^ As of 15^th^ June 2020 there were 867,000 cases nationally with São Paulo contributing a fifth of these.^6^ In March 2020, the Municipal Health Department of the municipality of São Caetano do Sul – part of the Greater Metropolitan Region of São Paulo – began to develop a clinical and testing platform to organize its COVID-19 response. The aim was to provide universal detection and management of symptomatic cases and their contacts. The platform was developed in partnership with two local universities – the Municipal University of São Caetano do Sul (USCS) and the University of Sao Paulo (USP) – and called “Corona São Caetano”.

Large scale community-based observational cohorts are difficult to establish under epidemic circumstances, particularly if the risk of exposure for research personnel is high. Hence, most COVID-19 epidemiological and clinical studies have been hospital-based,^7–9^ and therefore tend to include more severe cases whose findings may not be generalizable to the general population.^10^ The objectives of this study were to describe the epidemiological indicators of the early phase of the programme rollout; and to describe the clinical, virologic and natural history features (including hospitalization and deaths) of SARS-CoV-2 infection among patients identified in primary care.

## METHODS

### Setting

The municipality of São Caetano do Sul has a population of 161,000 inhabitants.^11^ The active aging index (i.e., the ratio of population aged >60 yr / population aged ≤14 yr) is 135, compared to the Brazilian average of 52, reflecting an aging population;^11^ its Human Development Index is one of the highest in the country; nearly all (97·4%) children aged 6-14 are in education and 31% of the population have completed higher education^12^ (Brazilian national average is 11%).

### Corona São Caetano platform

Residents of the municipality aged 12 years and older with suspected COVID-19 symptoms were encouraged to contact the dedicated Corona São Caetano platform via the website (access at https://coronasaocaetano.org/) or by phone. They were invited to complete an initial screening questionnaire that included socio-demographic data; information on symptoms type, onset and duration; and recent contacts.

Patients meeting the suspected COVID-19 case definition (i.e., having at least two of the following symptoms: fever, cough, sore throat, coryza, or change in/loss of smell (anosmia); or one of these symptoms plus at least two other symptoms consistent with COVID-19) were further evaluated, whilst people not meeting these criteria were reassured, advised to stay at home and contact the service again if they were to develop new symptoms or worsening of current ones. Patients were then called by a medical student to complete a risk assessment. All pregnant women, and patients meeting pre-defined triage criteria for severe disease (see Supplemental Material), were advised to attend a hospital service - either an emergency department or outpatient service, depending on availability. All other patients were offered a home visit for self-collection of a nasopharyngeal swab.

### Sample collection

Nasopharyngeal swabs (NPS – both nostrils and throat) were collected at the patients’ homes under the supervision of trained healthcare personnel. A link to a video (https://youtu.be/rWZzV2ZP7KY) was sent to the patients, before the home visit, to provide guidance on self-collection procedures. Healthcare personnel were instructed to maintain a distance of six feet from the patient and to wear personal protective equipment at all times. Samples were immediately put on a cool box between 2-8°C and stored at 4°C in a fridge until shipment to the lab within 24 hours.

### Follow-up procedures

Patients testing SARS-CoV-2 RT-PCR positive were followed up to 14 days (a maximum of 7 phone calls) from completion of their initial questionnaire. They were contacted every 48 hours by a medical student who completed another risk assessment and recorded any ongoing or new symptoms. Patients testing RT-PCR negative were followed up by the primary health care program for their residential area. They were advised to contact the platform for a new consultation if they developed new symptoms. Starting on May 19^th^, when serological testing became available, RT-PCR-negative patients were re-contacted to offer antibody (IgG/IgM combined) testing 14 days after their initial registration as long as they had become asymptomatic.

### Study dates

The Corona São Caetano programme was launched on 6th April 2020 and is still ongoing at the time of writing. For this analysis, we opted to include all patients making their first contact with the programme between 13th April and 13th May 2020. This comprises the first 31 days of the response, having excluded the first week, which corresponded to a pilot phase designed to test instruments before roll-out. The period of follow-up (last date of data extraction) was 4th June 2020, to account for the accrual period (three weeks) of possible hospitalizations in the last included patients.

### Laboratory methods

Due to shortages of some reagents, two RT-PCR platforms were used at different times during the study: ALTONA RealStar® SARS-CoV-2 RT-PCR Kit 1·0 (Hamburg, Germany) and the Mico BioMed RT-qPCR kit (Seongnam, South Korea). For serology we tested 10μL of serum or plasma (equivalent in performance) using a qualitative rapid chromatographic immunoassay (Wondfo Biotech Co., Guangzhou, China), that jointly detects anti-SARS-CoV-2 IgG/IgM. The assay has been found to have a sensitivity of 81·5% and specificity of 99·1% in a US study^13^. In our local validation, after two weeks of symptoms, the sensitivity in 59 RT-PCR confirmed cases was 94·9%, and specificity in 106 biobank samples from 2019 was 100%.

### Statistical methods

We estimated the contribution of our primary platform to COVID-19 diagnosis in São Caetano do Sul. We compared the number of cases diagnosed in our programme with official data released by the Municipal Department of Health in its daily bulletins (accessed here https://coronavirus.saocaetanodosul.sp.gov.br).

Clinical and demographic data were extracted directly from the Corona São Caetano information system, with the last export on 5^th^ June, to allow for follow-up of patients at the end of the study period. To analyse clinical presentation, we first calculated the proportion and exact binomial 95% confidence intervals (CI) of cases reporting each symptom in the three testing groups: SARS-CoV-2 RT-PCR positive; RT-PCR negative / seropositive; and RT-PCR negative / seronegative. We next combined RT-PCR and serology positive cases to make confirmed COVID-19 group, and those negative on both tests to make a SARS-CoV-2 negative control group. We express the association between each symptom and a positive COVID-19 diagnosis as odds ratios (OR) and 95% CIs. For RT-PCR-positive patients, we grouped the follow-up questionnaire responses into two-day intervals from symptom onset. In order to illustrate symptom trajectories through time, we calculated the proportion of questionnaire responses where a given symptom was present for each time window.

Next, we assessed associations between RT-PCR cycle thresholds (Cts) and other clinical features. ALTONA and MiCo BioMed RT-PCR kits each separately amplify two different SARS-CoV-2 viral genes, as such each patient had two Ct values. There was a high concordance between Cts for the two genes within each kit (Figure S4), and we opted therefore to use the mean of the two Ct values for each patient in all analyses. We calculated univariable associations between Cts and age, sex, delay from symptom onset to NPS collection, and presenting symptoms using simple linear regression. We then built a multivariable linear regression model to assess independent associations between presenting symptoms and RT-PCR Cts. As age, sex, and time of swab collection may confound this relationship we included these variables, as well as the RT-PCR platform (ALTONA vs MiCo BioMed), as covariates in the model.

For RT-PCR positive patients (followed up for 14 days), hospitalizations and deaths were extracted from the study platform. To extend the follow-up period and to capture RT-PCR negative patients and those initially triaged to hospital (no study follow-up), hospitalization and vital status was confirmed by linkage with two administrative databases: the municipal epidemiological surveillance dataset, as well as the state-wide influenza-like illness notification system (SIVEP-Gripe). Linkage was last performed on 5^th^ June 2020, 23 days after the last patient was enrolled. Categorical patient characteristics were compared according to hospitalization status using a Chi-squared or Fisher exact test. Continuous variables were compared using the Wilcoxon rank sum test.

The cohort sample included consecutive cases presenting to the Corona São Caetano program and a formal sample size calculation was not performed. Missing data were excluded. All analyses were conducted in R Software for Statistical Computing, version 3.6.3.^14^

### Ethics

The study was approved by the local ethics committee (Comissão de Ética para Análise de Projeto de Pesquisa - CAPPesq, protocol No. 13915, dated June 03, 2020). The committee waived the need for informed consent and allowed the development of an analytical dataset with no personal identification for the current analysis.

## RESULTS

### Epidemiological and programmatic indicators

Between 13^th^ April and 13^th^ May 2020, there were 2,073 presentations, from 2,011 individual patients, that met the criteria for a suspected COVID-19 case. At initial phone interview, 132 (6%) potential cases were advised to go directly to a health service based on the triage questions, and 12 (0·6%) because of pregnancy. Only four (3%) of referred patients were admitted to hospital and none died.

In total 1,583 individual patients were tested with RT-PCR for SARS-CoV-2; 444 (28·0%, 95%CI 25·9%-30·3%) were positive. The proportion of positive results was stable over the study (Figure S1). Among the RT-PCR negative group, 604 (53% of 1,136) underwent serology testing, of whom 52 (8·6%, 95%CI 6·6% - 11·1%) were seropositive. The median [IQR] time from symptom onset to serology collection was 31 [26 – 37] days. The age-sex structure of patients being tested differed from the underlying population of São Caetano do Sul (Figure S2) with an overrepresentation of working-age adults and women. At the beginning of programme role out, 75% of notified COVID-19 cases in São Caetano do Sul were diagnosed in outpatient or hospital services. Over the study period, adherence to the programme increased, and by May 13^th^, 2020, 78% of cases in the municipality were diagnosed within our programme.

Of 444 RT-PCR positive patients eligible for longitudinal follow-up, 326 (73%) had their final follow-up visit at least 14 days after their initial presentation. Of the seven possible follow-up questionnaires, 384 (86%) COVID-19 patients completed three or more, and 162 (36%) completed all seven.

### Participant characteristics

Patient characteristics are shown in Table 1. Although women were overrepresented in the cohort, there were proportionally more males in the RT-PCR positive and seropositive groups compared to the seronegative group. Of note, 55% of RT-PCR negative/seronegative patients had completed higher education compared to 35% RT-PCR-positive patients (p < 0·001, Chi-squared test). The median number of days from symptom onset to swab collection was 5.0 (interquartile range [IQR], 4·0-7·0) among RT-PCR positive patients and 6·0 (IQR, 4·0-8·3) among RT-PCR negative/seropositive patients (p = 0·06, Wilcoxon rank sum) (Figure S3). Chronic respiratory disease was less frequent in RT-PCR positive than dual-negative patients.

**Table 1.**
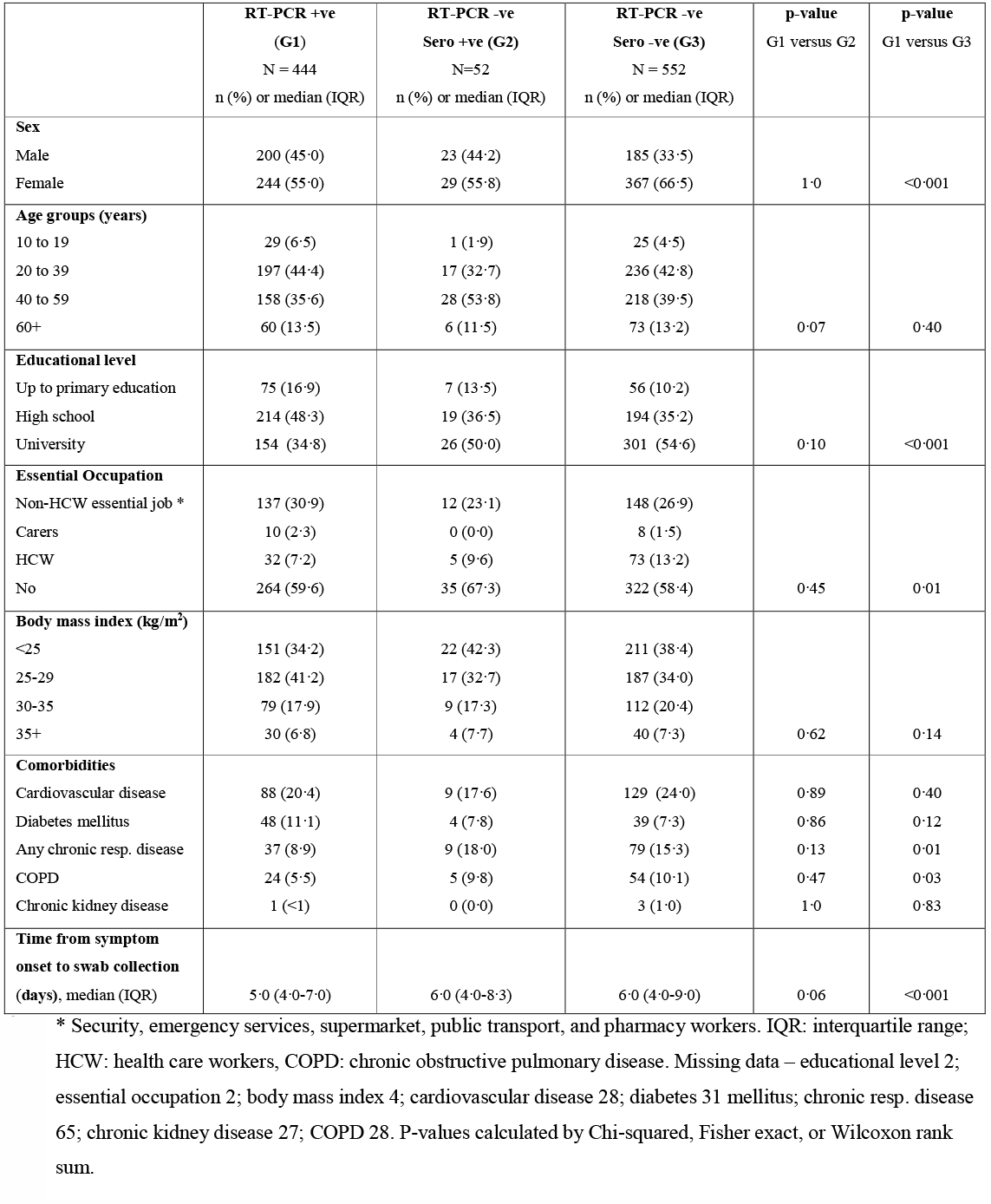
Demographic and clinical characteristics of 1,048 suspected COVID-19 cases undergoing diagnostic testing in the Corona São Caetano program

### Symptoms of COVID-19 at cohort presentation

The prevalence of individual symptoms at presentation is shown in Figure 2A stratified by final diagnostic category. The most frequent symptoms among RT-PCR and seropositive patients were headache (82% and 75%), myalgia (80% and 80%), cough (77% and 63%), and fatigue (77% and 79%) (Figure 3). Anosmia was present in 56% and 63% of RT-PCR positive and seropositive patients, respectively, compared to 30% in those testing doubly negative. A similar pattern was observed for ageusia (53% and 53% versus 30%).

**Figure 1.**
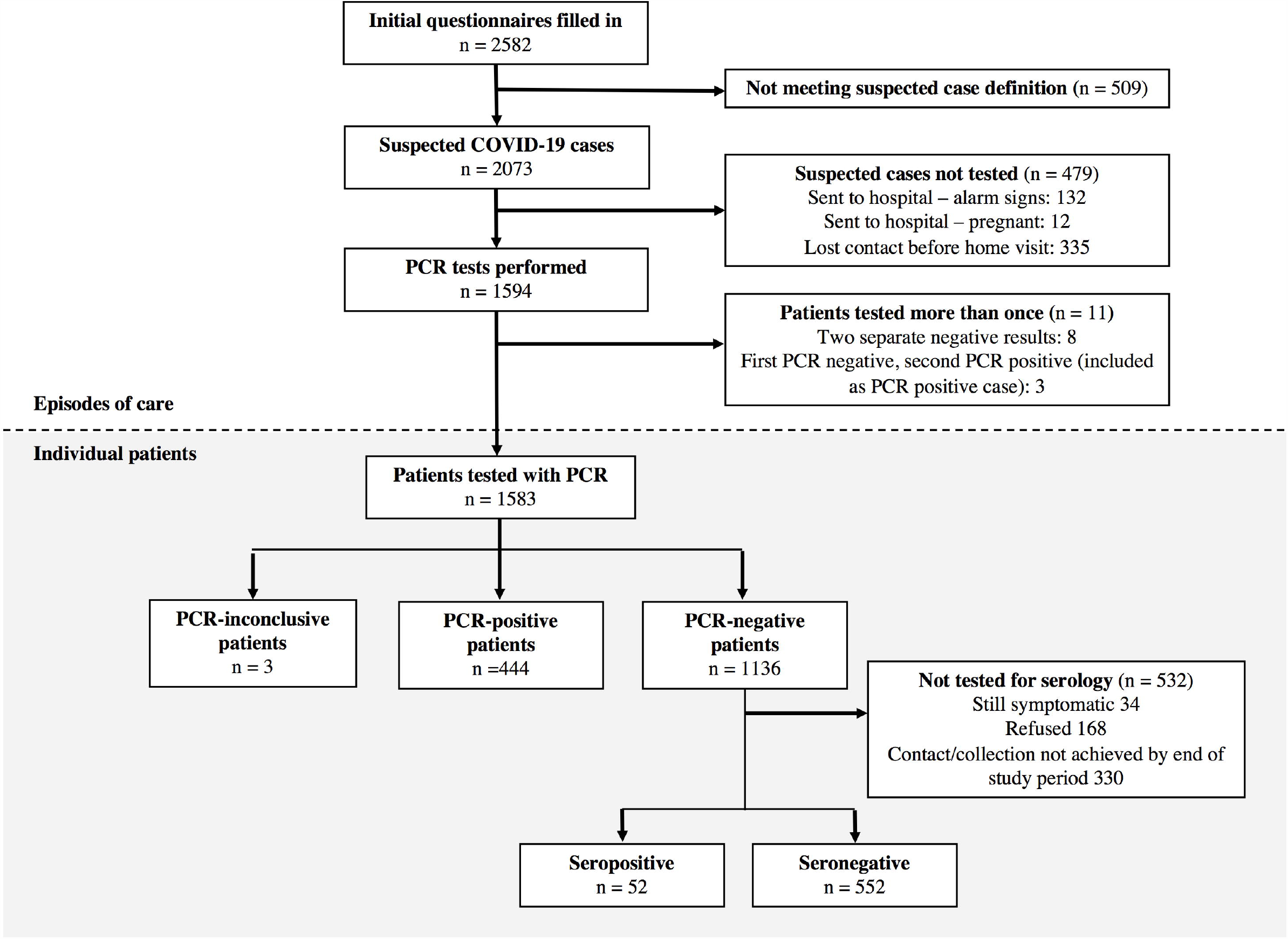
Patient flowchart for the Corona São Caetano platform between 13^th^ April and 13^th^ May 2020. In the upper section (white background) the numbers correspond to individual presentations to the system; among suspected cases 2,073 suspected cases, 60 had two presentations and one had three. In the lower section (grey background) numbers correspond to individual patients making up the final analytic groups.

**Figure 2.**
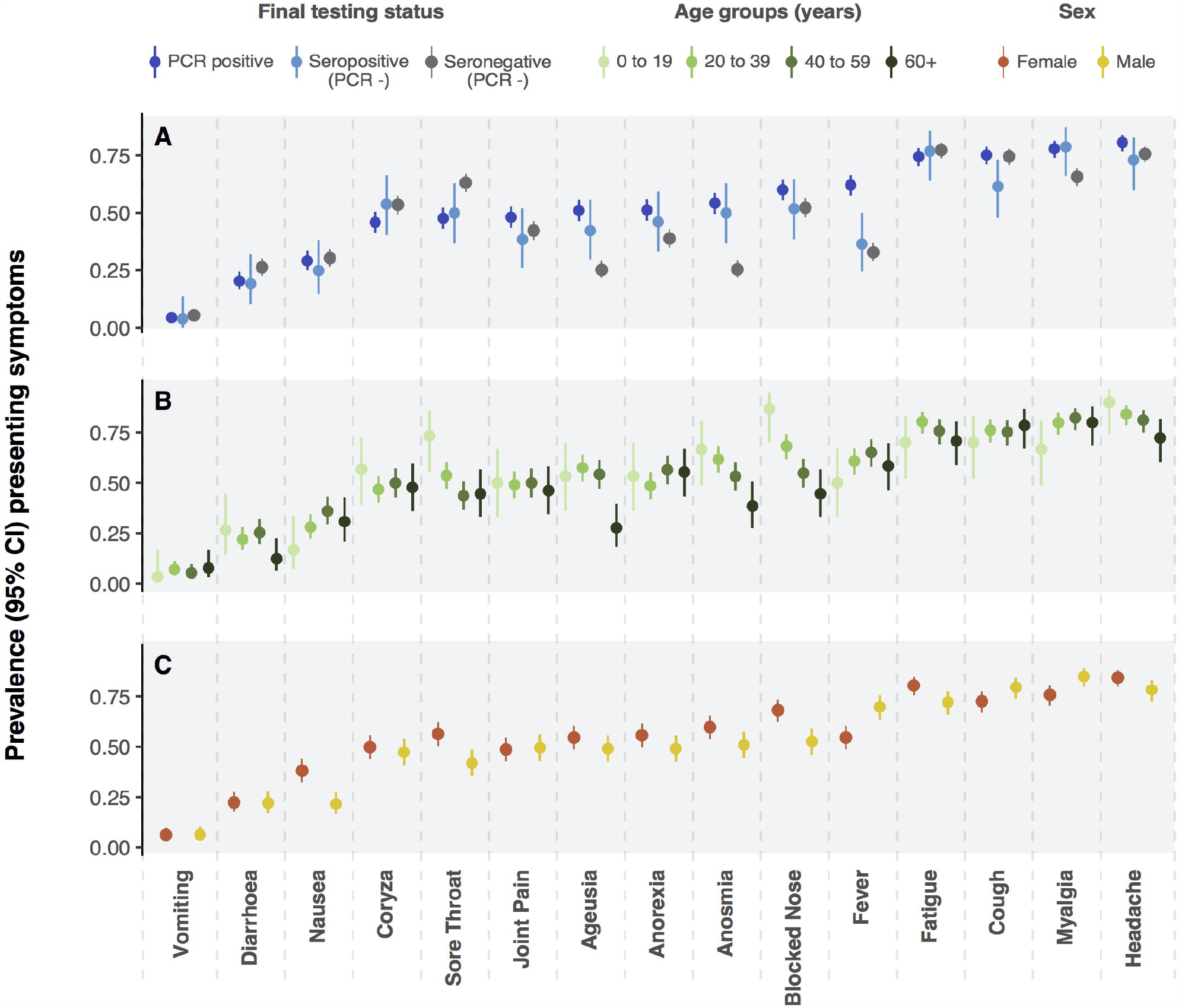
Panel A present prevalence (point) and exact binomial 95% confidence intervals (vertical lines) of symptoms at presentation among patients with suspected COVID-19 according to RT-PCR result and serostatus (A). Panels B and C present the prevalence of presenting symptoms among patients with COVID-19 (RT-PCR and serology positive) stratified by age (B) and sex (C).

**Figure 3.**
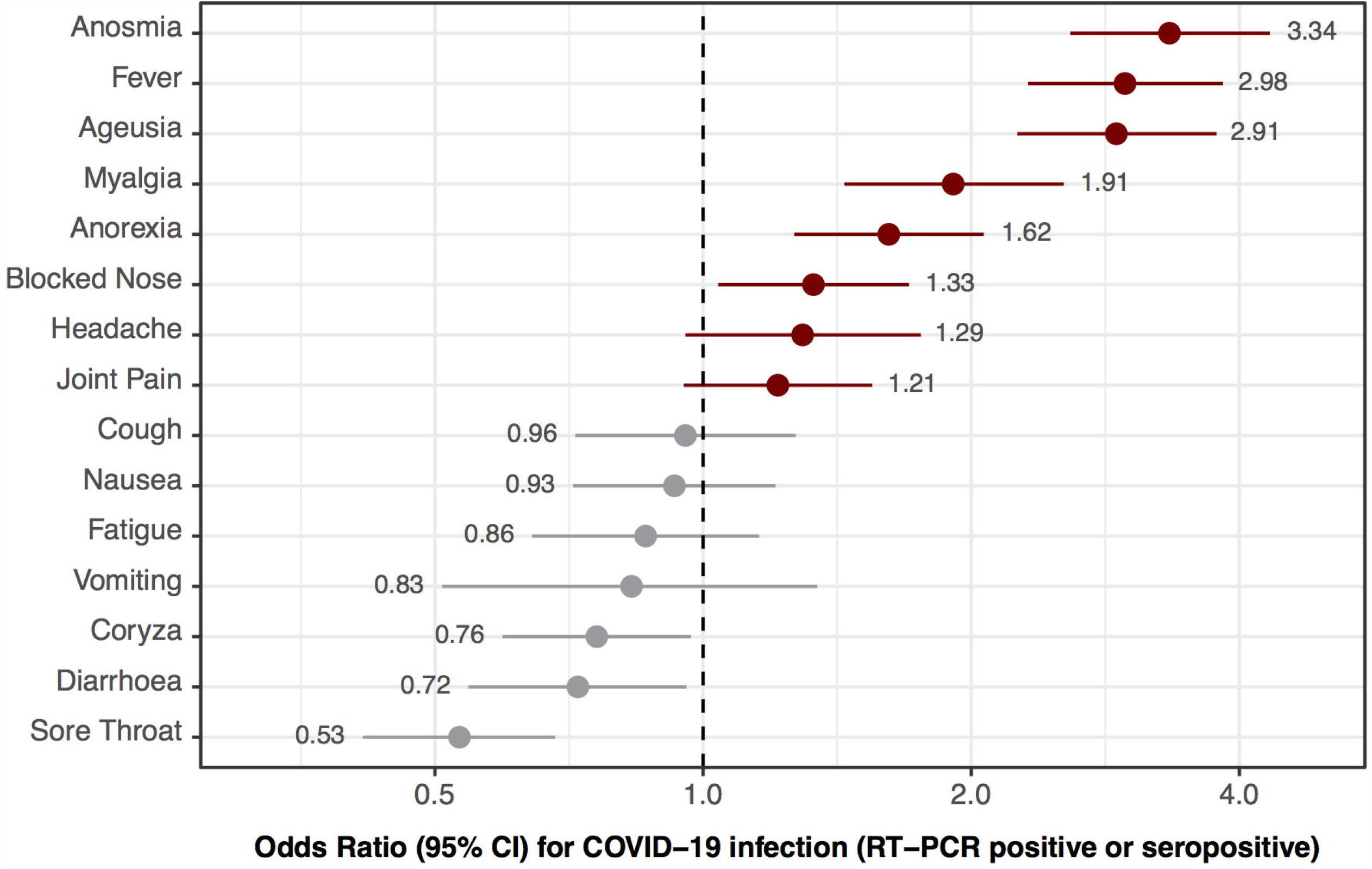
Odds ratios (black dot) and 95% confidence intervals (lines) for testing positive for COVID-19 (RT-PCR positive or serology positive) associated with the presence of each presenting symptom. Horizontal axis is on log scale. Point estimates of odds ratios are shown inline with their corresponding symptom.

The odds ratios for testing positive for SARS-CoV-2 (RT-PCR or serology) associated with each presenting symptom are shown in Figure 3. The symptoms with strongest associations were anosmia (OR 3·3, 95%CI 2·6-4·4), fever (3·0, 95%CI 2·4-3·9) and ageusia (2·9, 95%CI 2·3-3·8). The presence of sore throat (0·53, 95%CI 0·41-0·68) and diarrhoea (0·72, 95%CI 0·55-0·96) were associated with a negative SARS-CoV-2 test.

Among RT-PCR positive or seropositive patients, in general, younger patients presented with more symptoms, with mean [standard deviation] number of symptoms of 8·33 [1·92], 8·24 [2·39], 8·09 [2·46] and 7·05 [2·54], in those aged 12 to 19 years, 20 to 39 years, 40 to 59 years, and 60+ years, respectively (p= 0·008, Kruskal-Wallis test). In particular, upper respiratory tract symptoms - including coryza, blocked nose, ageusia, and anosmia - were more frequent in younger people (Figure 2B). The mean [sd] number of symptoms was greater in women (8·28 [2·41]) than men (7·72 [2·45]) (p = 0·005, Wilcoxon rank sum) (Figure 2C).

### Symptoms over time in SARS-CoV-2 RT-PCR positive patients

Figure 4 presents the symptom questionnaire responses over time for 444 RT-PCR positive patients. In general, constitutional symptoms – in particular fever, arthralgia, and myalgia – were prominent at symptom onset, with a large drop in the proportion reporting these symptoms after four to six days. By contrast, anosmia, and ageusia were most frequent among questionnaire responses at four to eight days and continued to be reported later in the illness course. Cough was highly prevalent at disease onset, with a third of questionnaires completed at 14 to 16 days positive for cough.

**Figure 4.**
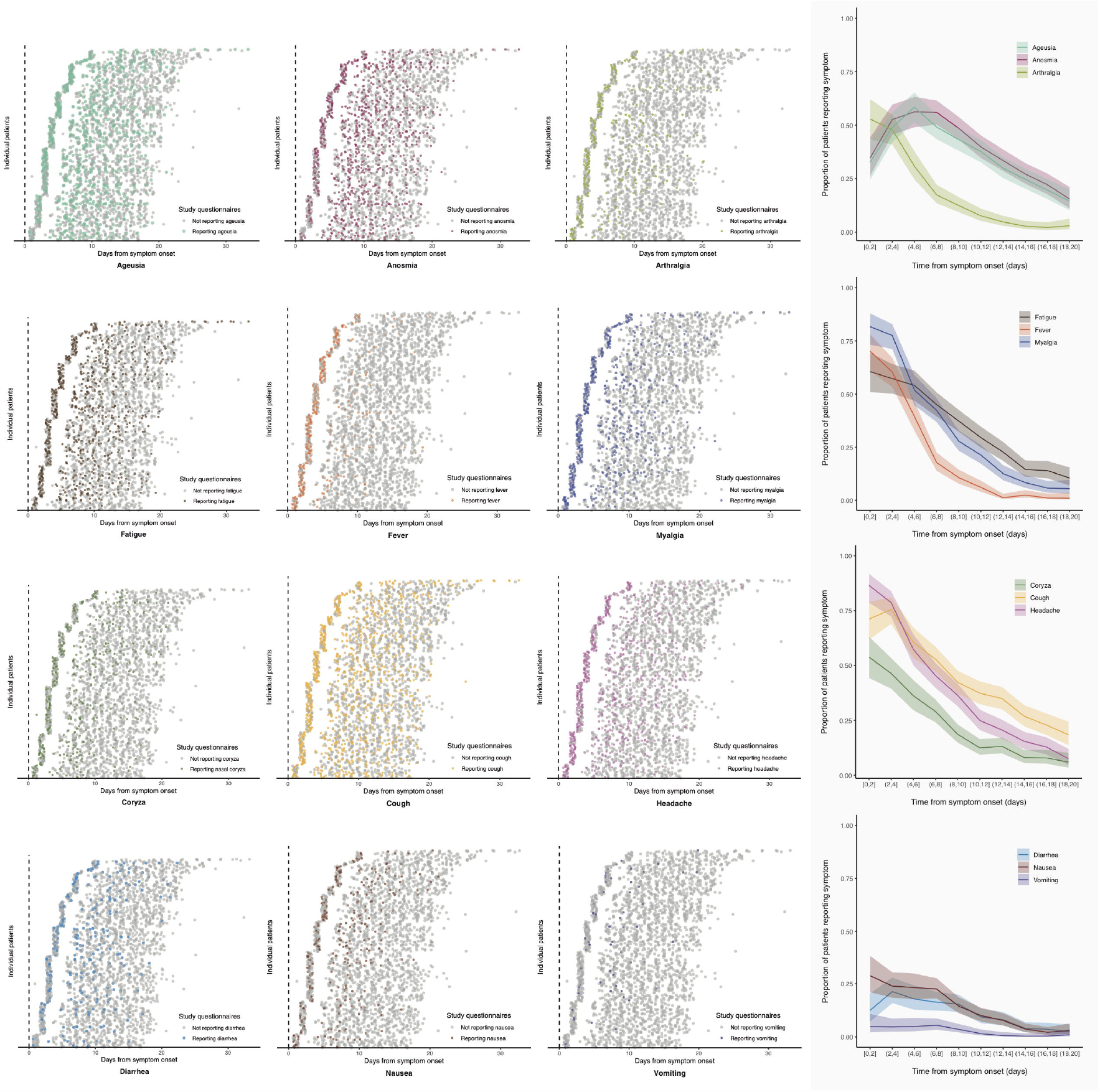
Left hand figures show symptoms at each follow-up questionnaire among patients testing RT-PCR positive and undergoing follow-up. Individual patients are stacked on the y-axis ordered according to the delay from symptom onset to presentation. Each point represents the response to a questionnaire and its position on the horizontal axis the time after symptom onset that the questionnaire was filled in. Grey points are questionnaires where the patient denied the presence of a given symptom. The coloured points correspond to questionnaires in which the patient reported a given symptom. The right-hand figures results from grouping the horizontal axis time into two-day windows and calculating the proportion of completed questionnaires in which each symptom was reported. The denominators for the horizontal axis groups (number of questionnaires completed within a given time window from symptom onset) are 104 at [0-2] days, 192 at (2-4], 185 at (4-6], 293 at (6-8], 338 at (8-10], 329 at (10-12], 335 at (12-14], 324 at (14-16], 280 at (16-18] and 201 at (18-20].

### Associations between SARS-CoV-2 RT-PCR Cycle threshold (Ct) values, and demographic and clinical features

Figure 5 shows the associations between mean RT-PCR cycle threshold and demographic features and symptoms at presentation. Older age was associated with lower cycle thresholds, with a change in mean Ct of −0·05 (95%CI −0·09 to −0·01) for each additional year of age. The mean difference in Ct value was −1·36 (95% CI −2·49 to −0·23) in men compared to women. For each doubling in the number of days from symptom onset to swab collection the mean Ct value increased by 3·28 (95%CI 2·33 to 4·03). Presenting symptoms of fever and arthralgia were associated with lower Cts, whereas anosmia, ageusia, vomiting, diarrhoea, and nausea were associated with higher Cts (Figure 6 and Table S1). After adjustment for age, sex, delay from symptom onset, and RT-PCR platform used, fever (−0·06, 95%CI −2·11 to −0·001) and arthralgia (−1·24, −2·18 to −0·10) remained associated with lower Cts, and anosmia (2·21, 1·0 to 3·29), ageusia (1·96, 0·88 to 3·0), and diarrhoea (1·36, 0·12 to 2·61) with higher Cts (Table S1).

**Figure 5.**
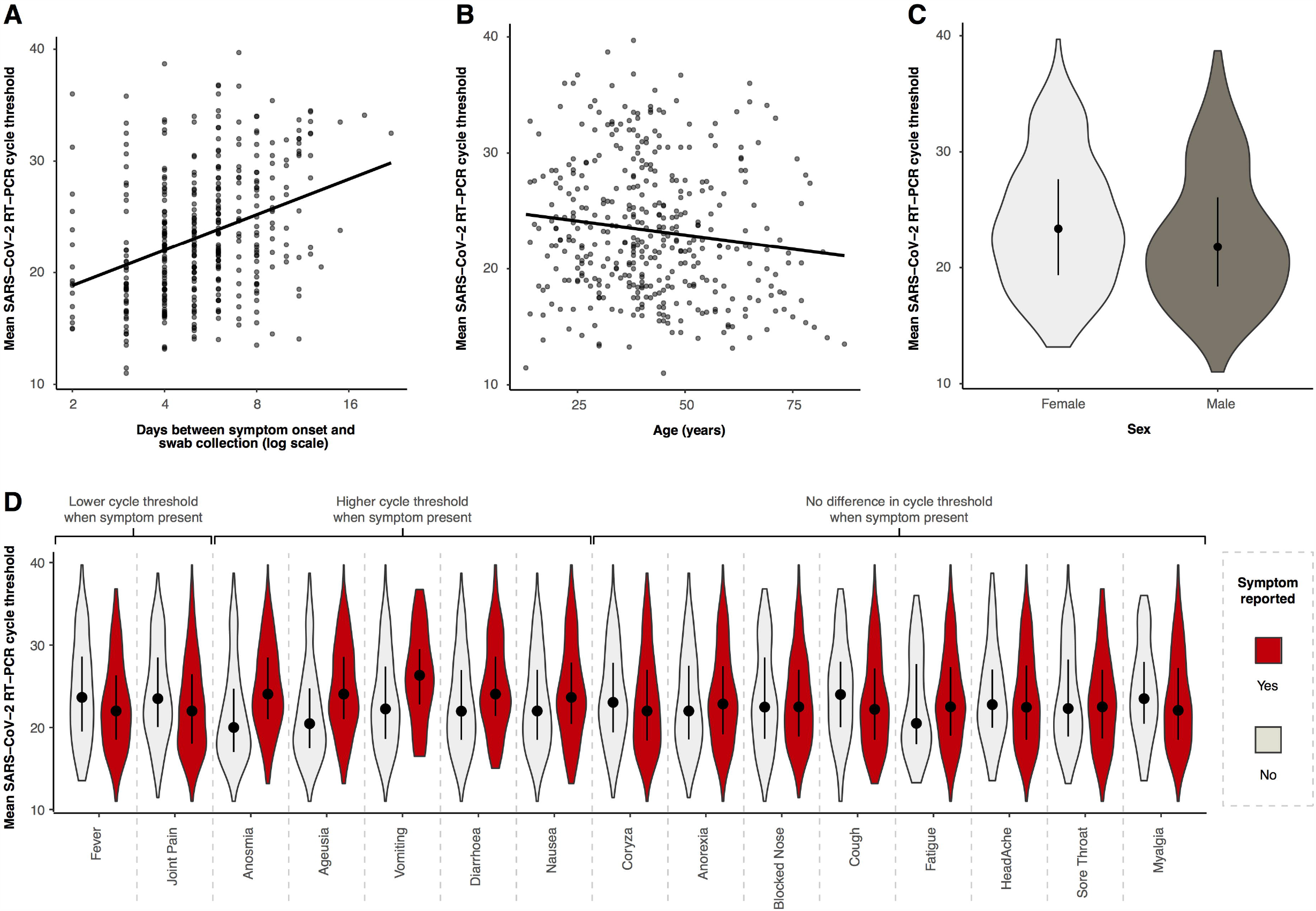
Relationship between mean RT-PCR cycle threshold (Ct) and day of illness course when the nasopharyngeal swab was collected (A), patient age (B), patient sex (C), and different symptoms at presentation. Panels A and B show the best fit linear regression lines, panels C and D are violin plots (rotated kernel density plots showing the full distribution of data) of the Ct values with median (black dot) and interquartile range (black line).

### Hospitalizations and deaths

Of the 444 RT-PCR positive patients, 30 (6·8%) had been hospitalized by 5^th^ June 2020, when the database linkage was last updated, and three (0·7%) had died; in-hospital mortality was therefore 10% (3/30). In 28 cases the date of admission was available. The median time from symptom onset to hospital admission was 7 (range 2 to 14) days. Among 1,136 RT-PCR-negative patients, six (0·5%) had been admitted to hospital. One (<0·01% of 1,136) of these six patients died. None of the 604 RT-PCR negative patients that underwent serology were admitted to hospital or died. Table 2 compares patient characteristics by hospitalization status. Notably, hospitalized patients were older, had more cardiovascular comorbidities and were more frequently obese.

**Table 2.**
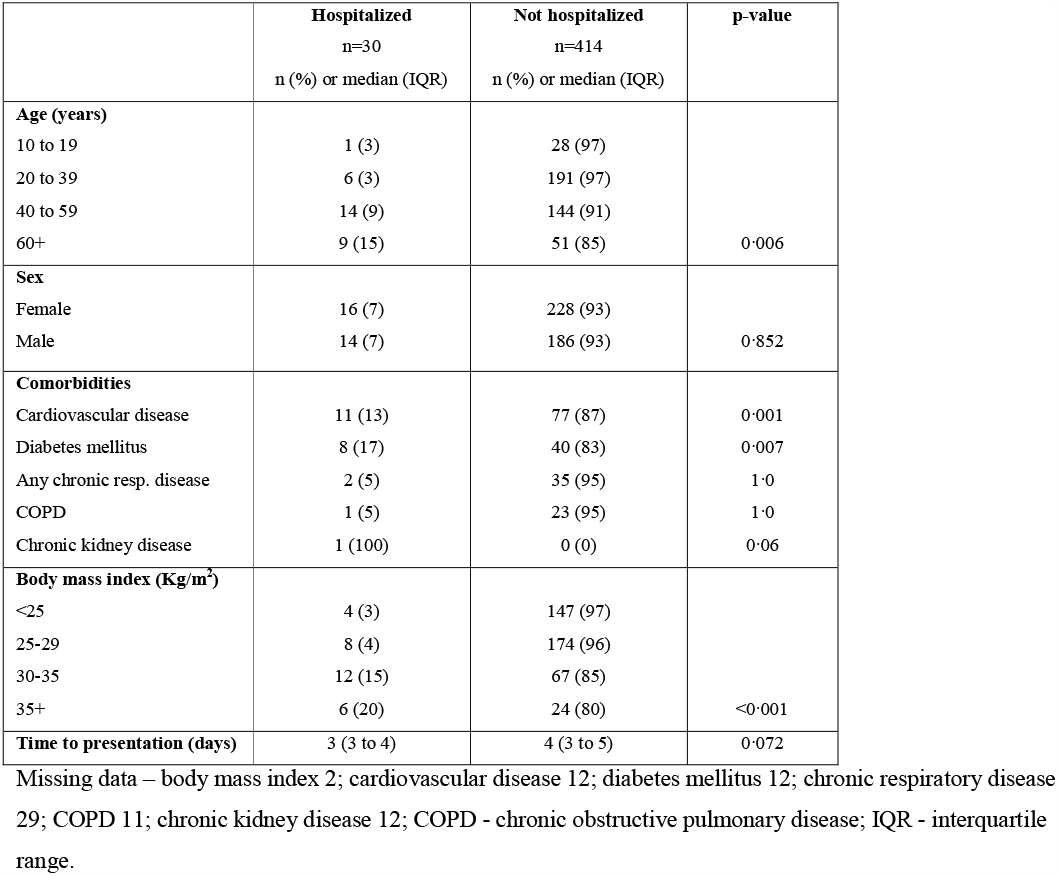
Characteristics of RT-PCR positive patients stratified by hospitalization status.

|  | **Hospitalized**<br>n=30<br>n (%) or median (IQR) | **Not hospitalized**<br>n=414<br>n (%) or median (IQR) | **p-value** |
| --- | --- | --- | --- |
| **Age (years)** |  |  |  |
| 10 to 19 | 1 (3) | 28 (97) |  |
| 20 to 39 | 6 (3) | 191 (97) |  |
| 40 to 59 | 14 (9) | 144 (91) |  |
| 60+ | 9 (15) | 51 (85) | 0.006 |
| **Sex** |  |  |  |
| Female | 16 (7) | 228 (93) |  |
| Male | 14 (7) | 186 (93) | 0.852 |
| **Comorbidities** |  |  |  |
| Cardiovascular disease | 11 (13) | 77 (87) | 0.001 |
| Diabetes mellitus | 8 (17) | 40 (83) | 0.007 |
| Any chronic resp. disease | 2 (5) | 35 (95) | 1.0 |
| COPD | 1 (5) | 23 (95) | 1.0 |
| Chronic kidney disease | 1 (100) | 0 (0) | 0.06 |
| **Body mass index (Kg/m<sup>2</sup>)** |  |  |  |
| <25 | 4 (3) | 147 (97) |  |
| 25-29 | 8 (4) | 174 (96) |  |
| 30-35 | 12 (15) | 67 (85) |  |
| 35+ | 6 (20) | 24 (80) | <0.001 |
| **Time to presentation (days)** | 3 (3 to 4) | 4 (3 to 5) | 0.072 |

Missing data – body mass index 2; cardiovascular disease 12; diabetes mellitus 12; chronic respiratory disease 29; COPD 11; chronic kidney disease 12; COPD - chronic obstructive pulmonary disease; IQR - interquartile range.

## DISCUSSION

We present a community-based cohort of suspected COVID-19 cases recruited through a primary care initiative in the Brazilian municipality of São Caetano do Sul. Offering RT-PCR testing to all patients presenting with symptoms compatible with COVID-19, the positivity rate was 28%, with 8·6% of those testing negative subsequently found to be seropositive - i.e. > 35% of the cohort had a diagnosis of COVID-19. Anosmia, ageusia, and self-reported fever provided the greatest diagnostic value in identifying COVID-19. The rate of hospitalization and deaths among RT-PCR positive patients was low, at 6·8% and 0·7%, respectively. Our results provide important information on the clinical presentation, diagnostic testing and natural history of COVID-19 identified in the community.

Extrapolating the seropositivity rate among RT-PCR negative patients to the 532 that were not tested with serology, we estimate that an additional 46 seropositive cases would have been identified. This corresponds to a false-negative rate of 18% among potential symptomatic COVID-19 cases. This is lower than a recent pooled analysis: nadir of 20% at three days post-symptom onset.^15^ Viral load peaks around the time of symptom onset and remains high over the first symptomatic week (also see Figure 5A).^16,17^ Consistent with this, we found a slightly longer delay to swab collection in RT-PCR false-negative patients than RT-PCR positive patients (Figure S4).

COVID-19 presents in a similar way to other respiratory viral illnesses. Indeed, in our cohort the most common symptoms of COVID-19 - such as cough, fatigue, headache, etc. - were reported with a similar frequency among patients testing negative. It is therefore important to have identified anosmia, ageusia, self-reported fever, myalgia, and anorexia as the symptoms with greatest value in the differential diagnosis of COVID-19 in primary care. Conversely, sore throat and diarrhoea - both considered symptoms of COVID-19 in other settings –^18^ were more frequently due to other aetiologies in this primary care context. These results are robust for a number of reasons. Firstly, our sample is representative of the population of interest - i.e. consecutive patients with suspected COVID-19 in the community - instead of extrapolating from hospital cases. Symptom data were collected prospectively, eliminating recall or interviewer bias. Finally, we have a control group of patients who were negative for both RT-PCR and serology, minimizing misclassification due to false negative RT-PCR. In our study, the proportion of patients with a positive SARS-CoV-2 RT-PCR requiring hospitalization was low (7%). Early reports from China were of 13·8% of cases being severe^19^, but this value was lower when under ascertainment of cases was accounted for.^20,21^ This is because our cohort reflects mild to moderate cases, as severely ill patients are likely to have attended hospital directly. As such, only 3% of patients we triaged to attend health services were ultimately hospitalized, possibly due to self-selection of patients presenting to our service. Supporting this notion, our overall case fatality ratio among RT-PCR positive patients was 0·7%.

Our study has some limitations. Serology was not performed on all RT-PCR negative patients due to on-going symptoms, loss to follow-up, or patient refusal. Of note, none of the RT-PCR-negative patients that were admitted to hospital underwent serology testing. This suggests that patients who were not tested with serology may have had a higher prevalence of COVID-19 than those that were tested. In addition, imperfect serology test performance (81% sensitivity)^13^ will introduced false-negative results. Taken together, these biases may have underestimated the true seroprevalence among RT-PCR-negative cases, as well as the false-negative rate of RT-PCR. The latter calculation may also have been influenced by the inclusion of RT-PCR positive patients in the denominator, introducing an incorporation bias.^22^

A key strength to our study relates to the provision of primary healthcare in Brazil and its symbiosis with medical training nationwide. Primary health care - within the family health strategy (*Estratégia Saúde da Família*) - is cantered around a healthcare unit with a multi-professional team that is responsible for all residents in the immediate catchment area ^23^. São Caetano do Sul has 100% coverage with the family health strategy, and medical students from the municipal university (USCS) are integrated into the healthcare teams and progressively trained from the first year of medical school. Our initiative took advantage of this existing system, with the addition of an online platform allowing remote clinical assessment and follow-up. The suspension of normal clinical training at the medical school provided the workforce. The partnership with the University of São Paulo, which provided the laboratory diagnostics, created the unique opportunity to establish our prospective community cohort of suspected and confirmed COVID-19 cases. But we believe that this infrastructure can be implemented in other regions with less resources, now understanding the key steps and problems in the implementation. The second phase of the platform is now focusing on contact tracing from index cases identified by molecular or serological testing, using a rapid response team and rapid serological testing.

As in most places around the globe, the Brazilian National Health System is underfunded. Nevertheless, the fact that primary health care infrastructure is well established in many areas in Brazil, would allow the rapid deployment of similar strategies, and at low cost. A primary healthcare approach to the COVID-19 pandemic using a bespoke computer platform and telehealth to control the activities has been paramount to properly delineate the characteristics and dynamics of the disease at community level and plan a multifaceted public health response accordingly. Other respiratory disease such as influenza, measles, or tuberculosis may benefit from similar infrastructure.

## Data Availability

Data may be made available to researchers following appropriate local ethical approval.

## AUTHOR CONTRIBUTIONS

FL, MC, SC, MC, RB, and ES conceived and designed the study. FL, RG, and JB provided clinical oversight and supervision of medical students. FL, MC, LB, HD, and SS collected and curated the data. MC, TM, LV, and LS performed the laboratory analysis. LB performed the formal statistical analysis with assistance from FL, SS, NA, PM, ES. LB, FL, PM and ES wrote the first draft, and all authors reviewed, contributed to and approved the final version.

## ACKNOWLEDGMENTS

The municipal health department of São Caetano do Sul (Secretaria Municipal de Saúde da Prefeitura de São Caetano do Sul) funded the establishment and implementation of the platform. We also acknowledge an award from FAPESP (2018/14389-0) and the UK Medical Research Council (MR/S0195/1) to the Brazil-UK Centre for Arbovirus Discovery, Diagnosis, Genomics and Epidemiology (CADDE).

## REFERENCES

1 World Health Organization. (2020). Pneumonia of unknown cause – China. (https://www.who.int/csr/don/05-january-2020-pneumonia-of-unkown-cause-china/en/, Accessed 2020-06-18).

2 World Health Organization. Critical preparedness, readiness and response actions for COVID-19 (Interim Guidance) (https://www.who.int/publications-detail/critical-preparedness-readiness-and-response-actions-for-covid-19, Accessed 15 June 2020).

3 Hellewell J, Abbott S, Gimma A, et al. Feasibility of controlling COVID-19 outbreaks by isolation of cases and contacts. Lancet Glob Health 2020; 8: e488–96.

4 World Health Organization. Regional Office for the Western Pacific. (2020). Role of primary care in the COVID-19 response. (https://apps.who.int/iris/handle/10665/331921, Accessed 15 June, 2020).

5 World Health Organization. Situation report 27 (2020). (https://www.who.int/emergencies/diseases/novel-coronavirus-2019/situation-reports, Accessed 15 June 2020).

6 Brazilian Ministry of Health. (https://covid.saude.gov.br/, Accessed 15 June 2020).

7 Docherty AB, Harrison EM, Green CA, et al. Features of 20 133 UK patients in hospital with covid-19 using the ISARIC WHO Clinical Characterisation Protocol: prospective observational cohort study. BMJ 2020; m1985.

8 Richardson S, Hirsch JS, Narasimhan M, et al. Presenting Characteristics, Comorbidities, and Outcomes Among 5700 Patients Hospitalized With COVID-19 in the New York City Area. JAMA 2020; 323: 2052.

9 Zhou F, Yu T, Du R, et al. Clinical course and risk factors for mortality of adult inpatients with COVID-19 in Wuhan, China: a retrospective cohort study. Lancet 2020; 395: 1054–62.

10 Rauh AL, Linder JA. Covid-19 care before, during, and beyond the hospital. BMJ 2020; m2035.

11 Instituto Brasileiro de Geografia e Estatística (IBGE). (2020). São Caetano do Sul / Panorama. (https://cidades.ibge.gov.br/brasil/sp/sao-caetano-do-sul/panorama, Accessed 20 June 2020).

12 Human Development Atlas Brazil. (2020). São Caetano do Sul (http://atlasbrasil.org.br/2013/pt/perfil_m/sao-caetano-do-sul_sp, Accessed 20 June 2020).

13 Whitman JD, Hiatt J, Mowery CT, et al. Test performance evaluation of SARS-CoV-2 serological assays. medRxiv 2020; 2020.04.25.20074856.

14 R Core Team. R: A language and environment for statistical computing. R Foundation for Statistical Computing, Vienna, Austria. (https://www.R-project.org/. 2019).

15 Kucirka LM, Lauer SA, Laeyendecker O, Boon D, Lessler J. Variation in False-Negative Rate of Reverse Transcriptase Polymerase Chain Reaction–Based SARS-CoV-2 Tests by Time Since Exposure. Ann Int Med 2020; M20–1495.

16 To KK-W, Tsang OT-Y, Leung W-S, et al. Temporal profiles of viral load in posterior oropharyngeal saliva samples and serum antibody responses during infection by SARS-CoV-2: an observational cohort study. Lancet Inf Dis 2020; 20: 565–74.

17 He X, Lau EHY, Wu P, et al. Temporal dynamics in viral shedding and transmissibility of COVID-19. Nat Med 2020; 26: 672–5.

18 Vetter P, Vu DL, L’Huillier AG, Schibler M, Kaiser L, Jacquerioz F. Clinical features of covid-19. JBMJ 2020; m1470.

19 T. Team, Vital Surveillances: The Epidemiological Characteristics of an Outbreak of 2019 Novel Coronavirus Diseases (COVID-19)-China. China CDC Weekly 2020; 2: 113–22.

20 Salje H, Tran Kiem C, Lefrancq N, et al. Estimating the burden of SARS-CoV-2 in France. Science 2020; eabc3517.

21 Verity R, Okell LC, Dorigatti I, et al. Estimates of the severity of coronavirus disease 2019: a model-based analysis. Lancet Infect Dis 2020; 20: 669–77.

22 Worster A, Carpenter C. Incorporation bias in studies of diagnostic tests: how to avoid being biased about bias. CJEM 2008; 10: 174–5.

23 Macinko J, Harris MJ. Brazil’s Family Health Strategy — Delivering Community-Based Primary Care in a Universal Health System. NEJM 2015; 372: 2177–81.

